# Replacing non-emergency bleeps and long-range pagers with a hospital-wide, EHR-integrated secure messaging system: an implementer report

**DOI:** 10.1101/2022.11.21.22282572

**Authors:** Ari Ercole, Claire Tolliday, William Gelson, James Rudd, Ewen Cameron, Afzal Chaudhry, Fiona Hamer, Justin Davies

## Abstract

**Introduction:** Obsolete bleep/long-range pager equipment remains firmly embedded in the NHS.

**Objective:** To introduce a secure, chart-integrated messaging system (Epic Secure Chat™) in a large NHS tertiary referral centre to replace non-emergency bleeps/long-range pagers.

**Methods:** The system was socialised in the months before go-live. Operational readiness was overseen by an implementation group with stakeholder engagement. Cutover was accompanied by a week of Secure Chat and bleeps running in parallel.

**Results:** Engagement due to socialisation was high with usage stabilising approximately 3 months after go-live. Contact centre internal call activity fell significantly after go-live. No significant patient safety concerns were reported.

**Discussion:** Staff engagement and uptake was excellent. The majority of those who previously carried bleeps were content to use personal devices for messaging because of user convenience after reassurance about privacy.

**Conclusion:** An integrated secure messaging system can replace non-emergency bleeps with beneficial impact on service.

## Introduction

In 2019, the UK Health and Social Care Secretary announced that the NHS should remove bleeps and pagers for non-emergency communication by the end of 2021 [1]. Whilst this technology is now in costly obsolescence and pilot studies have shown efficiency saving [2] using smartphone messaging, legacy equipment remains firmly embedded in the NHS.

Cambridge University Hospitals NHS Foundation Trust (CUH) uses a comprehensive Electronic Health Record (EHR-Epic systems corporation, Verona, USA) since 2014. An information-governance compliant messaging solution (Epic Secure Chat™) which allows for messaging from smartphones, tablets or from within the EHR itself (desktop). The system is fully integrated with the patient chart so that messages and all read/reply times become part of the patient record. The large-scale implementation of such an EHR-integrated messaging system to actively replace non-emergency bleeps/long-range pagers in a large NHS organisation has not been previously described.

## Setting

CUH is a large, tertiary referral centre in the East of England. With over 1,100 beds and approximately 16,000 NHS and honorary contract staff, it offers a diverse range of services. A significant EHR upgrade (from Epic 2017 to the November 2020 version) was undertaken during the implementation period bringing additional Secure Chat functionality. The implementation period also coincided with a major Wi-Fi infrastructure upgrade to give full coverage across the whole estate.

Our aim was to replace all bleeps/pagers apart from ‘cardiac arrest’, ‘major trauma’ and ‘fire’ with Secure Chat.

## Methods

Secure Chat was made available at our organisation in July 2021. A go-live date in early 2022 was initially chosen due to ongoing COVID-19 pandemic disruption and to leverage additional necessary Secure Chat functionality that would only become available after an Epic version upgrade planned for November 2021.

An implementation group with executive responsibility was formed with representation from the hospital’s divisional structure to oversee the project. Socialisation was achieved by a network of ‘clinical champions’ and through regular communications including trust bulletin items, face-to-face and online Q&A events as well as information on screensavers and posters and offering at-the-elbow support in clinical settings. An etiquette guide was published to define appropriate use of different methods of communication.

For safety a transition period where contact centre operatives would send messages both to Secure Chat and to existing bleeps for one week post go-live was planned. Secure Chat would not be available during (un)planned Epic outages for which the contingency was to fall back on an internal directory of alternative contacts securely maintained by the contact centre and this was widely publicised.

Secure Chat allows for various groups to enable team and role-based messaging. Because of system limitations at the time of the original implementation, our hospital had not fully implemented a sign-in system which we could leverage for automatic group creation. Instead, we created ‘opt-in’ groups to replicate existing roles, relying on staff to opt-in (out) at the beginning (end) of their duties.

Mean comparison was with t-tests; structural breaks were examined using the Chow test. Significance was taken at p<0.05.

## Results

### Technical

Secure Chat access was enabled for all members of staff with an Epic login (non-clinical users (such as contact centre agents) were not given security to view patient charts). Users who would previously have used bleeps were strongly encouraged to use their own personal devices although mobile phones (or pool phones) were provided in a relatively small number of cases where staff did not have a suitable device or were unwilling.

### Workflow

For the mobile app, onboarding involved installation of a CUH-specific profile and a website was set up for this. An initial manual batch activation step was subsequently automated using Blue Prism® robotic process automation software (Blue Prism group, Warrington, UK) so that registrations could be completed day and night.

The creation of opt-in groups was a major undertaking and had to be done centrally and no reliable list of baton bleep roles existed. An initial list of some 220 groups was compiled from information from clinical champions and existing bleep lists. After some local user acceptance testing, these groups were made available in December 2021. Inevitably creation, editing and deletion of groups was necessary, and this needed to be done centrally: a review process was set up to ensure consistency.

### Outcomes

Adoption through socialisation in the months before go-live across all staff groups was rapid (supplementary figure S1) across all staff groups with pharmacy (and pharmacy technicians) proving to be an unexpected early adopter. The original 4^th^ May 2022 go-live date was pushed back at a final go/no-go meeting to 8^th^ June 2022 due to isolated specialty-specific readiness concerns. Gross total messages sent plateaued at over 600,000 by 3 months after cutover. Opt-in group maintenance peaked before go-live (supplementary figure S2) although a significant maintenance burden occurred after the original 4^th^ May date.

Internal call data handled by contact centre operatives is shown in Figure 1. The average number of internal calls handled by contact centre operative fell from 720 to 614/day (p<0.0001) after implementation. Whilst average time/call increased marginally from 37 to 38 seconds (p=0.014), the total call duration/day fell overall by nearly an hour from 7.4 to 6.5 hours/day (p<0.0001). There was evidence of significant structural breaks for call numbers and average call time, but not for overall call time (p=0.01, 0.0003 and 0.06 respectively).

**Figure 1.**
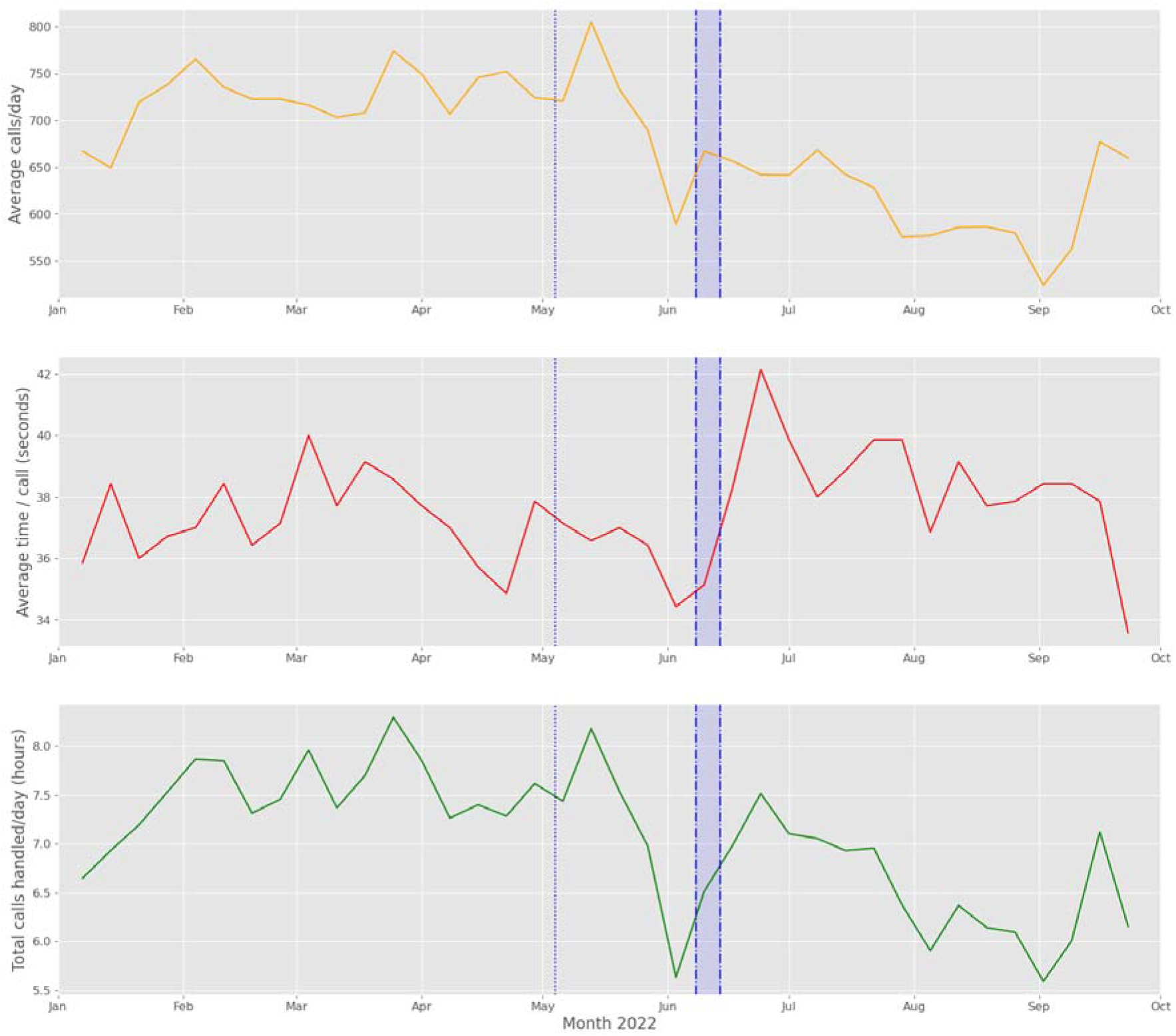
Uptake and organizational impact of Secure Chat implementation. **Top panel:** internal calls handled / day **Middle panel:** average contact centre time spent/call (seconds) **Bottom panel:** total call time (hours). Dotted line represents date of delayed initial go-live. Data is averaged by week to remove fluctuations from weekends.

Even without physically retiring the bleep system, call counts dropped from approximately 3,015 in the go-live week to 636 by middle of June 2022. A snapshot audit of the most frequently messaged devices suggested that they were not, in fact, largely no longer being actively answered.

## Discussion

Despite concerns that contact centre activity would increase with staff needing help regarding who was on call / who to message, we in fact found that internal calls fell significantly representing a reduction in agent workload. No significant risk events attributable to the Secure Chat implementation were reported on our incident reporting system which was monitored.

Although a minority of staff expressed reservations in the runup to go-live citing privacy concerns we were able to provide assurances; broadly speaking the vast majority of our staff were ultimately content to use their personal devices which offered substantial convenience advantages to users. The largest complaint received from users concerned inappropriate use of Secure Chat for non-urgent messaging: the etiquette guide setting out clear expectations was key central to empowering staff to challenge inappropriate messaging.

The beneficial reduction of internal call demand on our Contact Centre was unexpected but pleasing. A number of short (1-2hour) routine Epic upgrade outages have subsequently taken place (scheduled at weekends and night-time) during which time Secure Chat was not available. Concerns that the Contact Centre could be overwhelmed at these times have not materialised.

## Conclusions

We were able to effectively replace non-emergency bleeps/long-range pagers with a messaging system integrated with the patient chart in a large NHS academic hospital by the soft approach of socialisation before cutover. Discounting the time before our EHR upgrade in November 2021, we were able to do this in seven months with message numbers and support needs stabilising within approximately 3 months of go-live using existing infrastructure and without significant incident.

## Data Availability

Not applicable

## Data availability statement

Not applicable.

## Ethics statements

Not applicable.

## Competing interests

None at the time of implementation however AC will be leaving the role of Director of Digital at Cambridge University Hospitals in February 2023 to take up a post with Epic Systems Corporation.

## Acknowledgements

The authors acknowledge the support and engagement of the CUH eHospital team and management executive with this initiative. Special thanks are due to Joseph Scott, Richard Wallis, Benjamin Styllianou and Rose Cormie for their help in obtaining implementation data.

## Footnotes

### Contributors

All authors were involved in the implementation of the project. AE wrote up the project with approval from all other authors.

### Funding

Not externally funded.

### Competing interests

None declared.

### Provenance and peer review

Not commissioned; externally peer reviewed.

## SUPPLEMENTARY MATERIAL

**Figure S1.**
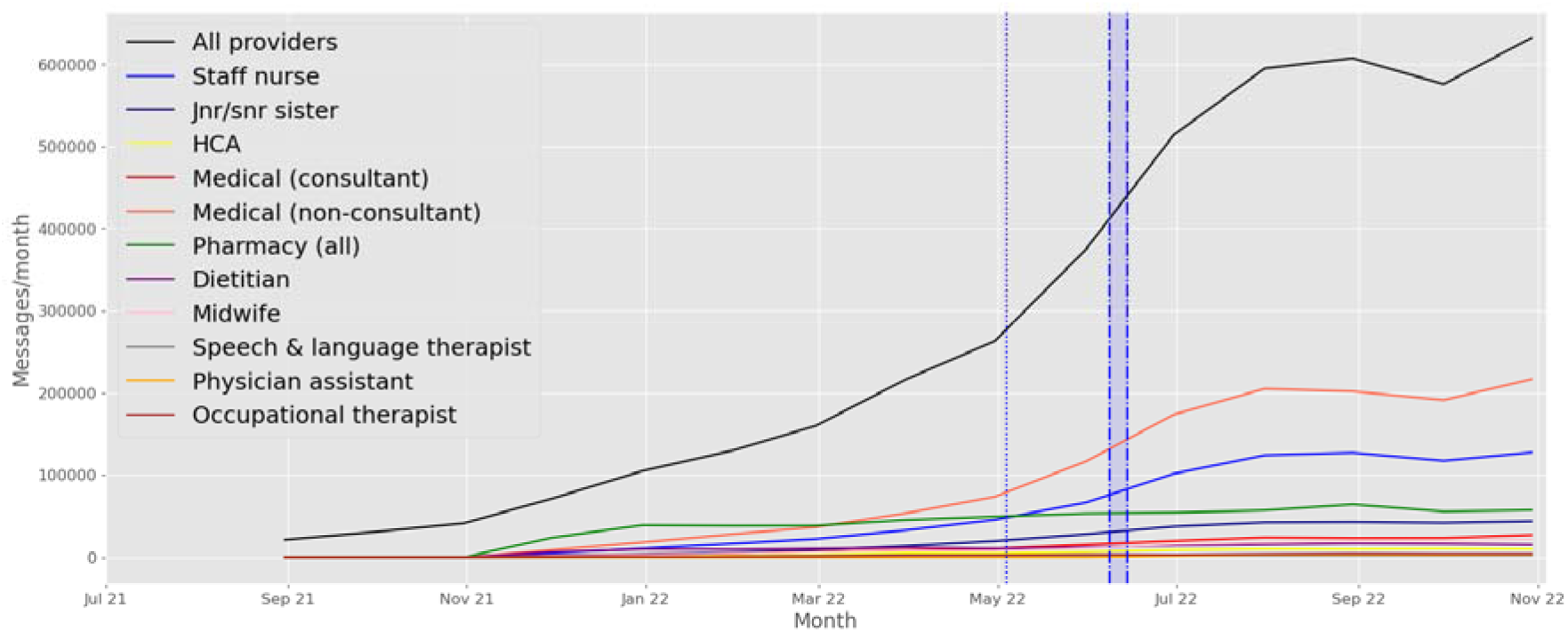
Gross messages per month, total for the organisation and by selected provider types). A plateau seems to be apparent some 3 months post-implementation.

**Figure S2.**
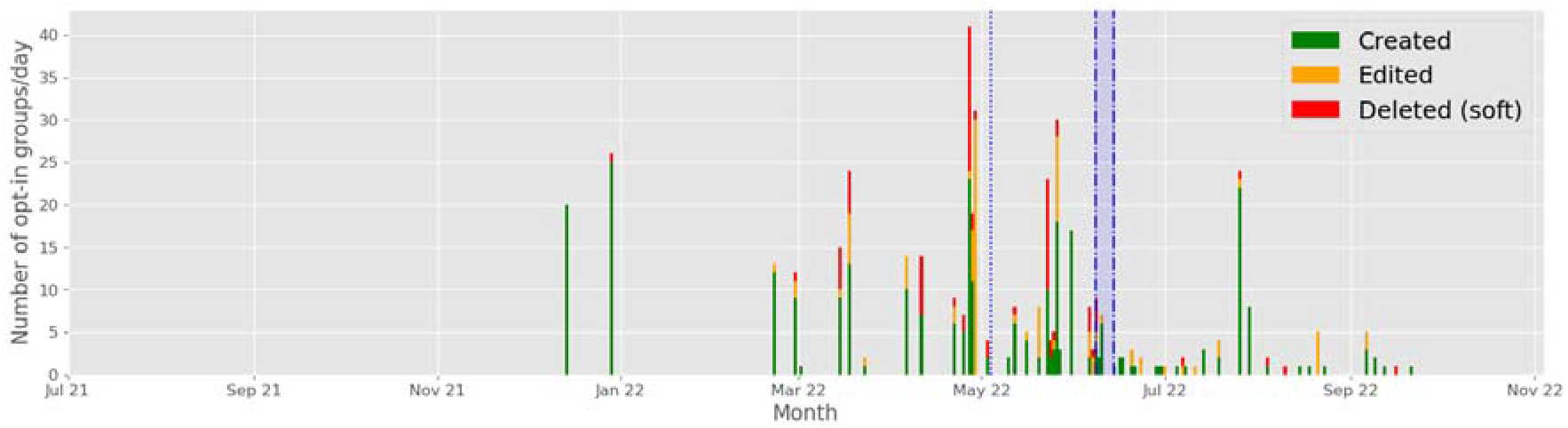
Opt-in group maintenance activity as a proxy for technical maintenance demand. Shaded blue area represents post-go live parallel bleep/secure chat period. Dotted line represents date of delayed initial go-live. Most of the activity occurred before go-live although spiked after the initial aborted date suggesting that some areas had not properly considered this at that time.

## Notes

### Funding Statement

This study did not receive any funding

